# Study on SARS-COV-2 transmission and the effects of control measures in China

**DOI:** 10.1101/2020.02.16.20023770

**Authors:** Bo Zhang, Hongwei Zhou, Fang Zhou

## Abstract

**Objective:** To reconstruct the transmission trajectory of SARS-COV-2 and analyze the effects of control measures in China.

**Methods:** Python 3.7.1 was used to write a SEIR class to model the epidemic procedure and a back propagation class to estimate the initial true infected number. The epidemic area in China was divided into three parts, Wuhan city, Hubei province (except Wuhan) and China (except Hubei) based on the different transmission pattern. A limitation factor for the medical resource was imposed to model the infected but not quarantined. Credible data source from Baidu Qianxi were used to assess the number of infected cases migrated from Wuhan to other areas.

**Results:** Basic reproduction number, R_0_, was 3.6 in the very early stage. The true infected number was 4508 in our model in Wuhan before January 22, 2020. By January 22 2020, it was estimated that 1764 infected cases migrated from Wuhan to other cities in Hubei province. Effective reproductive number, R, gradually decreased from 3.6 (Wuhan, stage 1), 3.4 (Hubei except Wuhan, stage 1) and 3.3 (China except Hubei, stage 1) to 0.67 (Wuhan, stage 4), 0.83 (Hubei except Wuhan, stage 2) and 0.63 (China except Hubei, stage 2), respectively. Especially after January 23, 2020 when Wuhan City was closed, the infected number showed a turning point in Wuhan. By early April, there would be 42073, 21342 and 13384 infected cases in Wuhan, Hubei (except Wuhan) and China (except Hubei) respectively, and there would be 2179, 633 and 107 death in Wuhan, Hubei (except Wuhan) and China (except Hubei) respectively.

**Conclusion:** A series of control measures in China have effectively prevented the spread of COVID-19, and the epidemic will end in early April.

## Introduction

On December 31, 2019, Hubei Provincial Health Commission announced the discovery of some clusters of pneumonia without definite cause [1]. Sequences of 2 complete viral genomes of 29.8 kilobases (HKU-SZ-002a and HKU-SZ-005b) obtained from 2 patients with the pneumonia were identified, and a novel lineage B coronavirus closely related to bat SARS-like bat-SL-CoVZXC21 (NCBI accession number MG772934) and bat-SL-CoVZC45 (NCBI accession number MG772933) was revealed [1]. World Health Organization (WHO) temporarily named the novel coronavirus as 2019-nCoV, and recommended that the novel coronavirus infected pneumonia be named 2019-nCoV acute respiratory disease. On Feb 11, 2020, WHO announced that 2019-nCoV infection has finally been given an official name COVID-19, and the Coronavirus Study Group (CSG) of the International Committee on Taxonomy of Viruses had named 2019-nCoV severe acute respiratory syndrome-related coronavirus 2, or SARS-CoV-2 [2]. It is believed that human infection with SARS-COV-2 first occurred in Wuhan on November 9, 2019 (95% credible interval: 25 September 2019 and 19 December 2019) [3]. 27 cases were initially reported, and 41 cases were reported by January 11, 2020, including 7 severe cases and 1 death [1].

SARS-COV-2 infected patients typically clinically manifested as fever, respiratory symptoms (cough, breathing difficulties); radiographic ground-glass lung changes; normal or below-average white blood cell, lymphocytes and platelet counts; hypoxemia; and liver and kidney dysfunction. It is reported that 5 / 41 cases can be combined with viral heart damage [4], and SARS-COV-2 infected patients can have gastrointestinal symptoms [1,5].

Some of the initial cases are geographically related to the Huanan Seafood Wholesale Market [6]. On January 24, 2020, Chan JF et al. reported the existence of human-to-human transmission of SARS-COV-2 [1]. The initial 425 reported cases included a total of 16 cases in 5 clusters as of mid-December, and some cases were found to have no apparent history of exposure [6]. In addition, a case of asymptomatic contact infection was reported by German researchers, first confirming that the latent cases can be contagious [7].

In the early stage of COVID-19 outbreak, the mean double time was estimated to be 7.4 days (95% confidence interval: 4.2 to 14); the basic reproductive number R_0_ was estimated to be 2.2 (95% confidence interval: 1.4 to 3.9); the mean incubation period was estimated to be 5.2 days (95% confidence interval: 4.1 to 7.0); and the mean serial interval was estimated to be 7.5 ± 3.4 days (95% confidence interval: 5.3 to 19) [6]. By January 29, 2020, the mortality rate of COVID-19 is about 2.1%, similar to the overall figures currently available [8].

The spread of infectious diseases is affected by various practical situations. When epidemic prevention measures (e.g. isolation, blockades, etc.) are taken, infectious diseases cannot spread under ideal circumstances. The effective reproductive number, R, refers to the number of secondary cases infected by one infected case during the actual spread of an infectious disease. In the absence of control measures, R=R_0_x, where x refers to the proportion of susceptible people. In the process of an infectious disease transmission, R will be reduced due to the implementation of special control measures and the reduction of the number of susceptible population [9]. After the outbreak of COVID-19, the Chinese government and people moved quickly to take measures to control the sources of infection and block the routes of transmission, and the spread of the epidemic was effectively curbed. Meanwhile the lack of proper diagnosis tools, the focus on the more severe cases or the overcrowding of hospitals make the reporting rate may be significantly lower in Wuhan city rather than elsewhere. In this study, mainly base on the Susceptible-Exposed-Infected-Removed (SEIR) model, we have studied the transmission dynamics of COVID-2019 under various practical situations. In this study, we divided the epidemic area into three parts, imposed the limitation factor of medical resource to model the infected but not quarantined especially in the early stage, and used a back propagation model to estimate the number of potential infections which is much larger than the official number announced. We strived to reconstruct the transmission trajectory of SARS-COV-2 in China and analyze the effects of China’s control measures.

## Materials and Methods

Our methodology was mainly based on the Susceptible-Exposed-Infected-Removed (SEIR) model, which is a very popular method where there is a considerable post-infection incubation period. The epidemic area was divided into three parts, Wuhan, Hubei (except Wuhan) and China (except Hubei) based on the different transmission pattern. A limitation factor for the medical resource was imposed to model the infected but not quarantined especially in the early stage. A back propagation model was used to estimate the number of potential infections which is much larger than the official number announced.

Python 3.7.1 was used to write a SEIR class to model the epidemic procedure and a back propagation class to estimate the initial true infected number.

### Susceptible-Exposed-Infected-Removed model

The classical SEIR models the number of people in the four state: susceptible (S), exposed (E), infected (I) and removed (R). The parameter beta controls how fast people move from S to E, and it is the product k and b where k stands for an average number of people of one infected person contact per day and b stands for the probability of infection. The k has a big impact to the number of infected and government takes various measures to cut down the parameter k. The sigma controls the speed from state E to state I, and gamma for state I to state R. The dynamic procedure of epidemic can be described by the following equation:

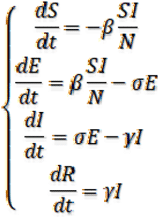

In our model we replaced the **σE** by max**(*θ, σE*)** where **θ** we call it limitation factor that models the max capability of medical resource that can be isolated timely. In the early stage, we assumed a very low limitation factor and many infected people could not be isolated timely or diagnosed and still infect others.

### back propagation model

The model is highly dependent on the initial values of S, E, I, R and other parameters, and to get better parameters is the key to have better results. We know that the true E group number is often been underestimated especially in the early stage. In this study, we used a back propagation method and the population out of Wuhan to estimate the true number of E group. The January 23, 2020 is a big milestone in this outbreak and government takes no major control actions before that day in Wuhan, and about 5 million people left Wuhan according to government press conference and a more concrete and credible data source from Baidu (https://qianxi.baidu.com/) shows that about 400 thousand people migrated into 8 provinces from Wuhan. We marked the 400 thousand number as M. Although there have been several reports of incubation time, there is no exact conclusion. And the diagnosis time may be about 3 days later than the onset time. We assumed that incubation period could be 7 days and 3 days more needed to diagnosis which means the duration from E to I could be 10 days. We collected 1568 infected number of these 8 provinces between January 23, 2020 to January 30, 2020 and then marked as m because of an average incubation of 7 days.

Baidu data shows that about 90% population left Wuhan flowed into other cities in Hubei, so the other cities in Hubei is the major areas except Wuhan. We use the same back propagation method above but with different parameters to get the initial true infected number of Hubei (except Wuhan).

## Results

### Early stage

We could estimate the true infected number in Wuhan at January 22, 2020 might be 4508 by the formula of 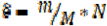, where N is the total population of Wuhan (11 million in 2018 statistics), which is much larger than the official announced 425. Then we could estimate the basic reproduction number (R_0_) could be 3.6 in the very early stage without control measures.

### Outcomes about Wuhan

The epidemic started from Wuhan and showed four significantly different stages (Fig 1). Stage 1 starts from December 8, 2019 to January 22, 2020. The true infected number was much more than official announced and could be 4508 in our model. These infected people were not treated effectively and isolated, which led to rapid spread of the epidemic. In the second stage, Wuhan was completely closed from January 23, 2020. The infected number growth curve had changed from exponential to nearly linear. In the third stage, the infected number showed a turning point and started to decline gradually. The last stage started from February 13, 2020, the infected number declined rapidly. Finally, the epidemic ended in early April, and there would be 42073 (95% confidence interval, 41673 to 42475) infected and 2179 (95% confidence interval, 2088 to 2270) death in Wuhan.

**Fig. 1.**
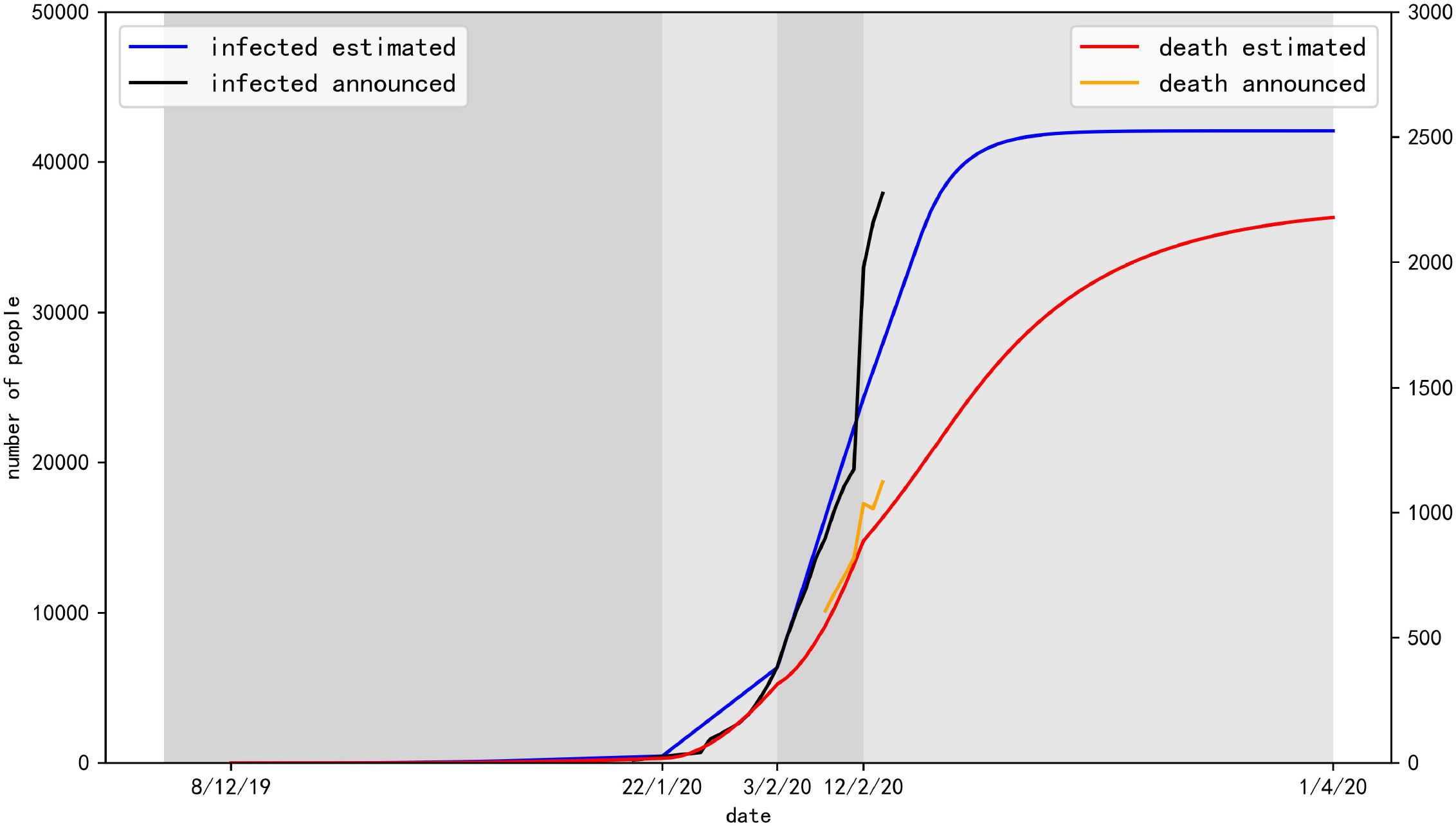
the predict number of infected and death in Wuhan in the four stages.

Key parameters and results in Wuhan prediction model were shown in table 1.

**Table 1.**
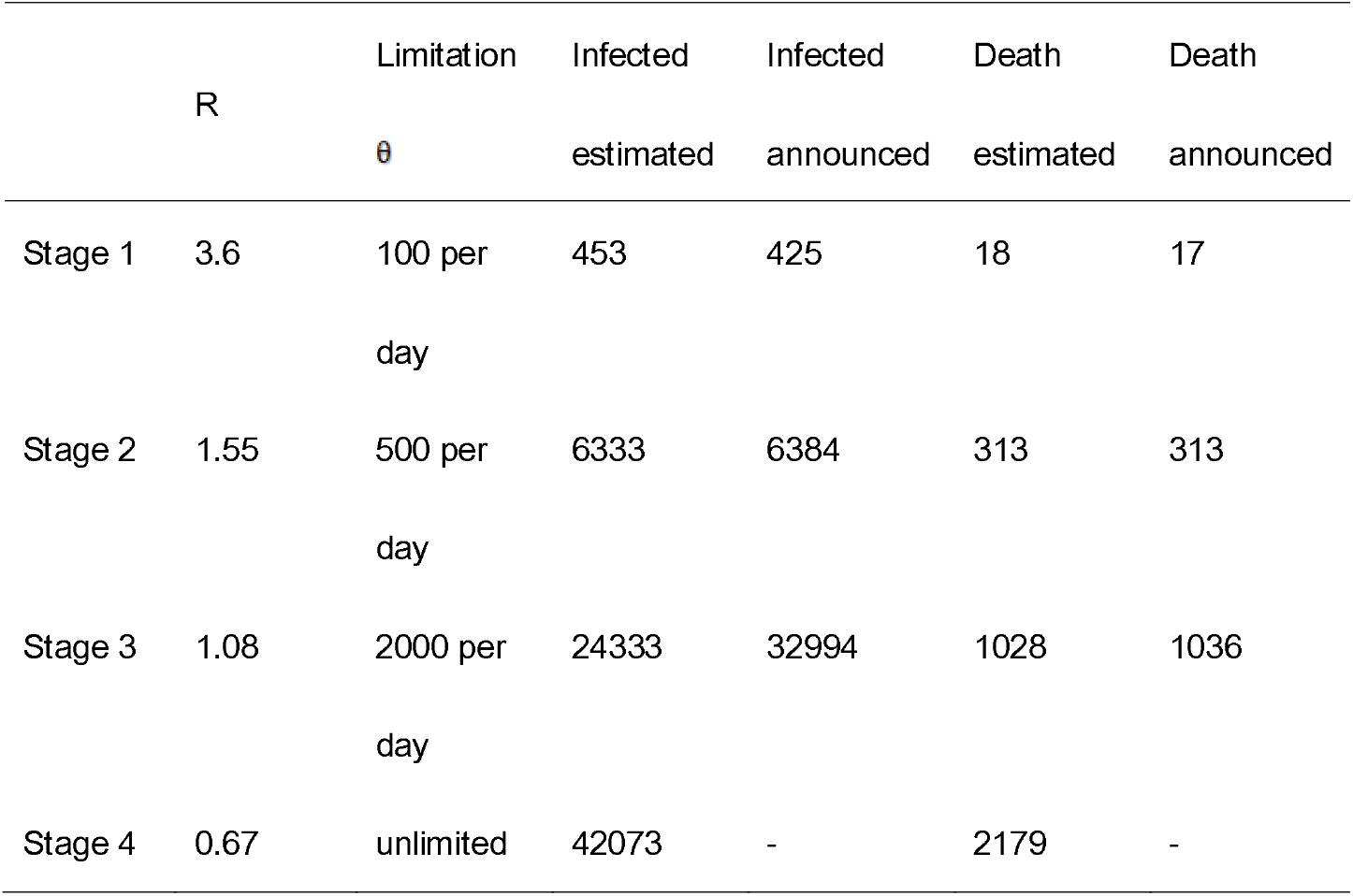
Key parameters and results in Wuhan prediction model

| | R | Limitation<br>$\theta$ | Infected<br>estimated | Infected<br>announced | Death<br>estimated | Death<br>announced |
| --- | --- | --- | --- | --- | --- | --- |
| Stage 1 | 3.6 | 100 per<br>day | 453 | 425 | 18 | 17 |
| Stage 2 | 1.55 | 500 per<br>day | 6333 | 6384 | 313 | 313 |
| Stage 3 | 1.08 | 2000 per<br>day | 24333 | 32994 | 1028 | 1036 |
| Stage 4 | 0.67 | unlimited | 42073 | - | 2179 | - |

### Outcomes about Hubei (except Wuhan)

The epidemic in Hubei (except Wuhan) showed three significantly different stages (Fig 2). According to the official announcement, about 5 million people left Wuhan from January 10, 2020 to January 22 2020, it means about 4.5 million people migrated into Hubei and about 1764 infected among them we estimated. In stage 1, the infected number increased exponentially. In stage 2, the added infected number reached the peak. In stage 3, the added infected number decreased significantly. Finally, there would be 21342 (95% confidence interval, 21057 to 21629) infected and 633 (95% confidence interval, 585 to 683) death in Hubei (except Wuhan).

**Fig. 2.**
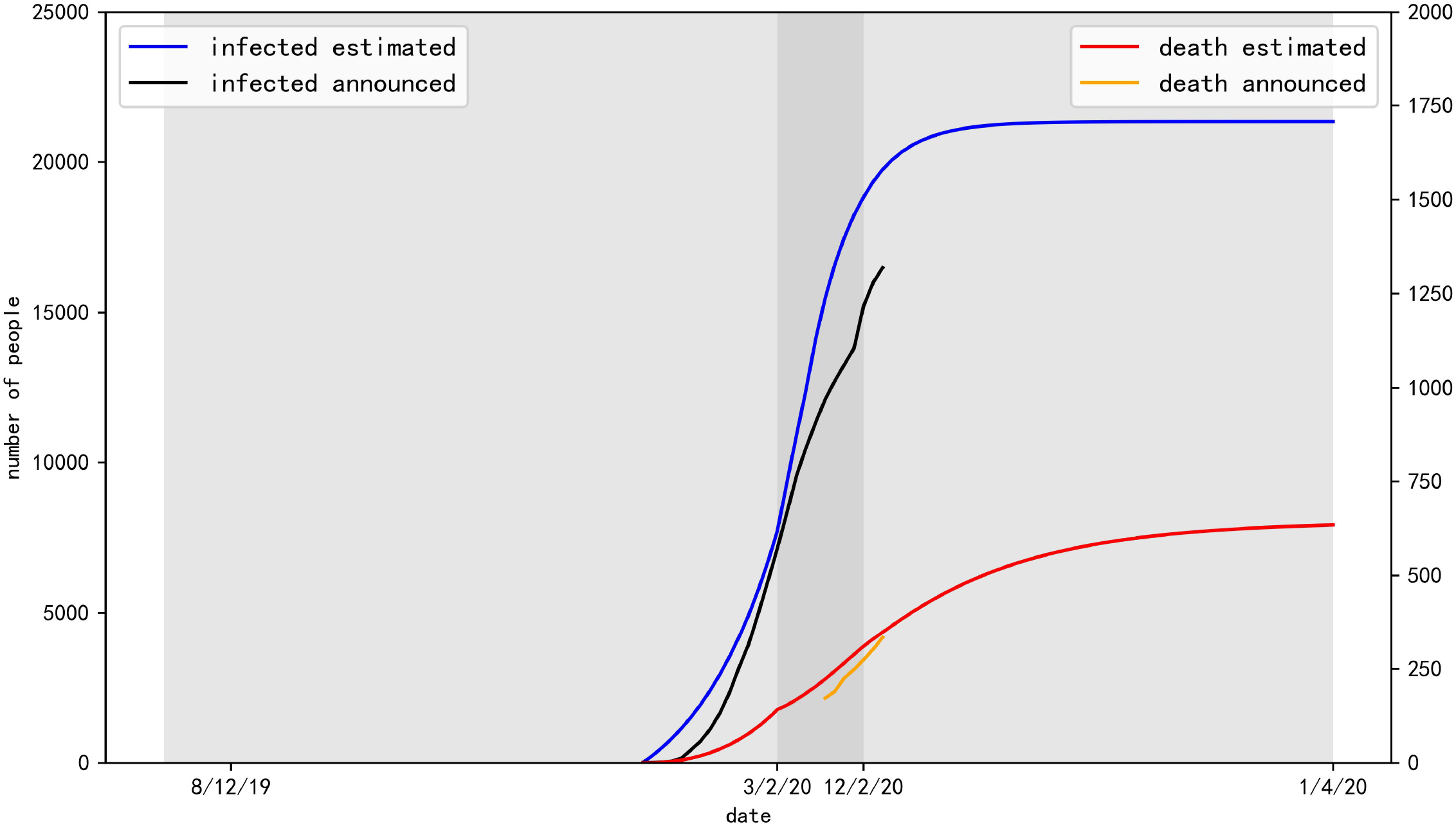
the predict number of infected and death in Hubei (except Wuhan) in the three stages.

Key parameters and results in Hubei (except Wuhan) prediction model were shown in table 2.

**Table 2.**
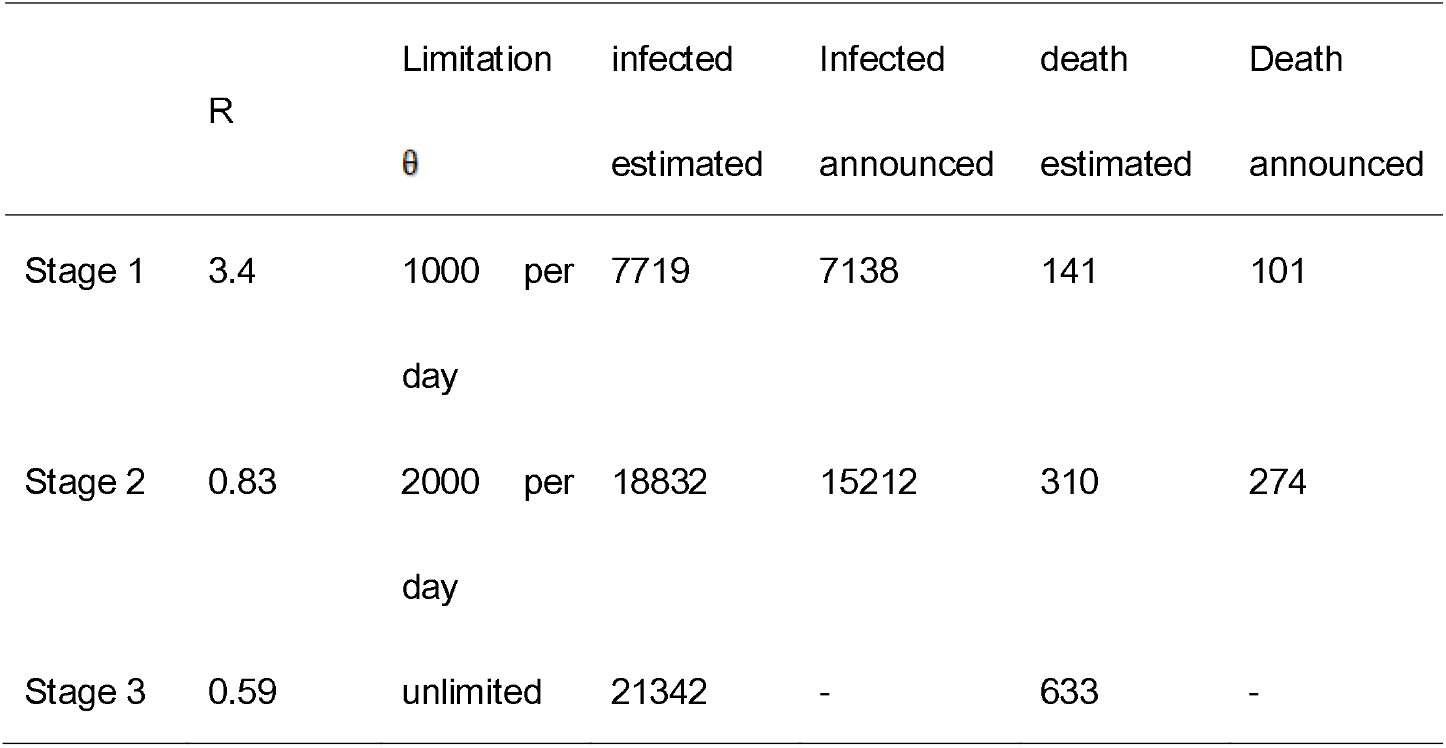
Key parameters and results in Hubei (except Wuhan) prediction model

| | R | Limitation<br>$\theta$ | infected<br>estimated | Infected<br>announced | death<br>estimated | Death<br>announced |
| --- | --- | --- | --- | --- | --- | --- |
| Stage 1 | 3.4 | 1000 per day | 7719 | 7138 | 141 | 101 |
| Stage 2 | 0.83 | 2000 per day | 18832 | 15212 | 310 | 274 |
| Stage 3 | 0.59 | unlimited | 21342 | - | 633 | - |

### Outcomes about China (except Hubei)

The epidemic transmission process could be divided into two stages (Fig. 3). In Stage one (from January 23, 2020 to February 3, 2020), the infected number increased rapidly. In stage two (from February 4), the infected number got its peek around February 3, 2020. It was estimated that the total infected and death number in China (except Hubei) could be 13384 (95% confidence interval, 13158 to 13612) and 107 (95% confidence interval, 87 to 128).

**Fig. 3.**
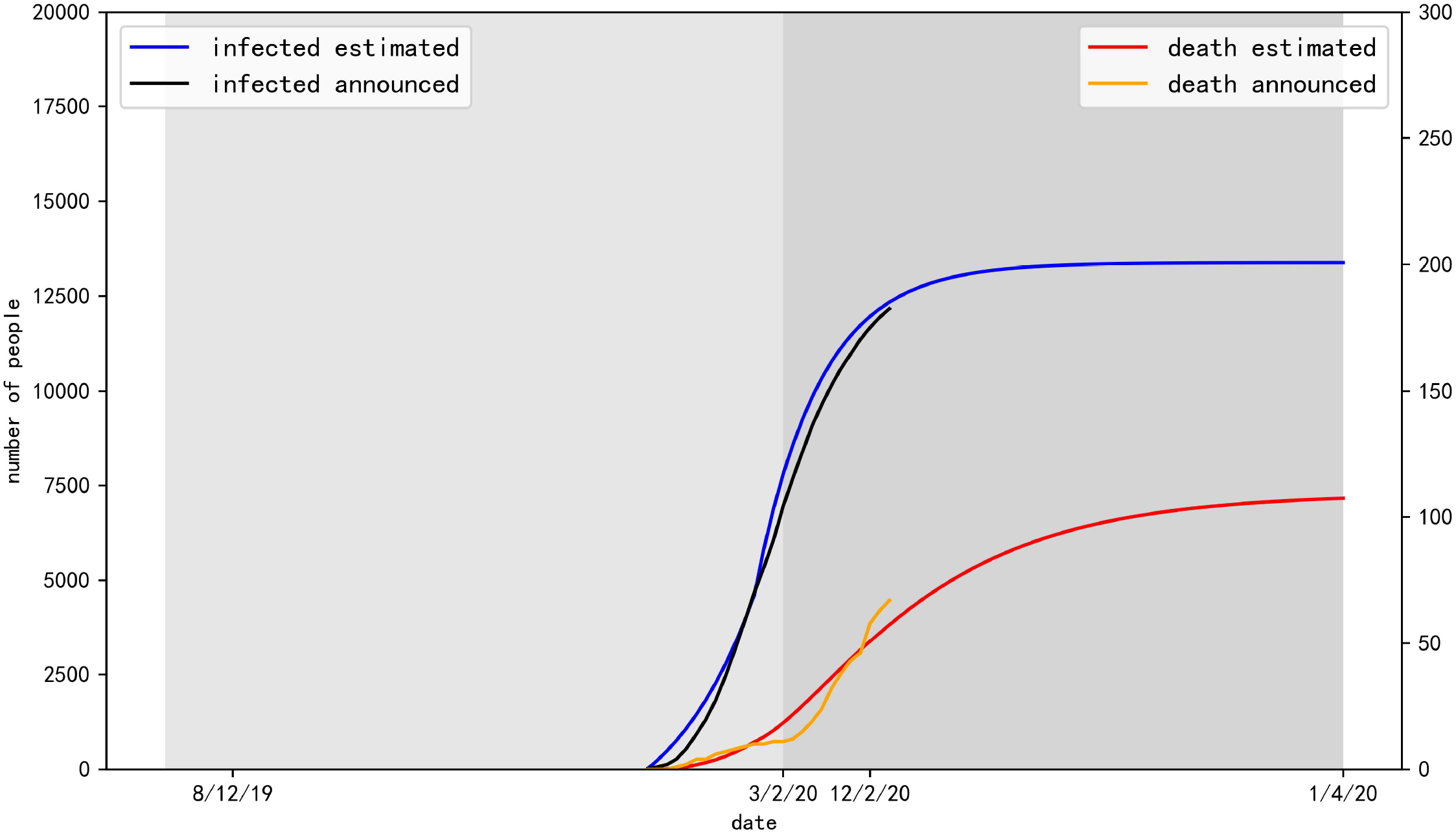
the predict number of infected and death in China (except Hubei) in the two stages.

Key parameters and results in China (except Hubei) prediction model were shown in table 3.

**Table 3.**
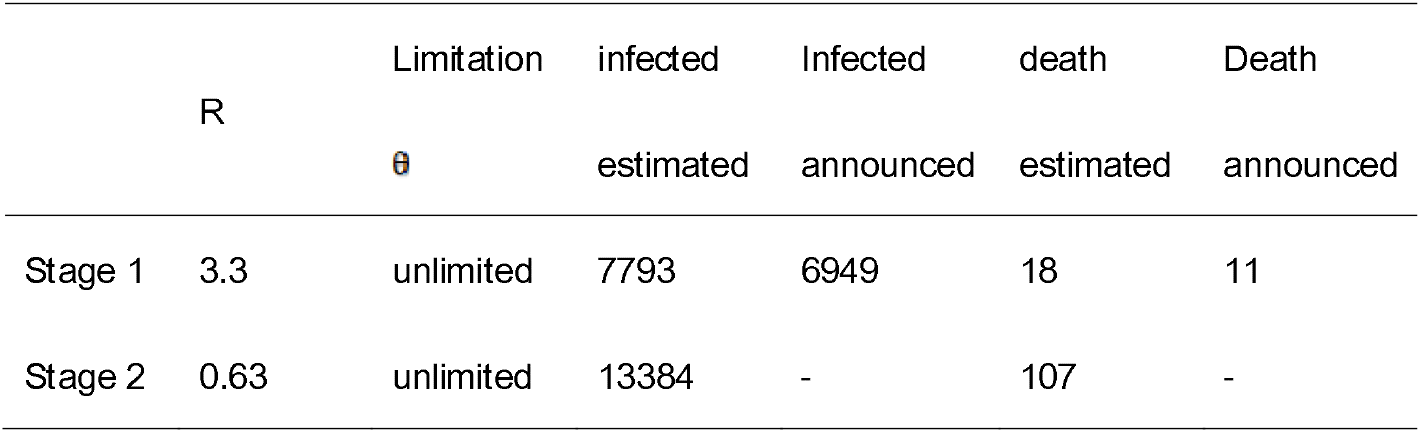
Key parameters and results in China (except Hubei) prediction model

## Discussion

Although COVID-19 also has gastrointestinal symptom and viral heart damage, it mainly causes SARS-COV-2 infected pneumonia [10]. SARS-COV-2 infected patients have reported respiratory illness with fever, cough and shortness of breath, and there is no vaccine to prevent COVID-19 [10]. American Centers for Disease Control and Prevention (CDC) combined epidemiologic risks (Travel from Wuhan City, close contact with a person who is under investigation for COVID-19 while that person was ill or close contact with an ill laboratory confirmed COVID-19 patient in the last 14 days before illness onset) and clinical features (fever, symptoms of lower respiratory illness) to diagnose suspected COVID-19 [10]. The definite diagnosis of COVID-19 can be make if the 344bp RNA-dependent RNA poly-merase (RdRp) gene related to severe acute respiratory syndrome (SARS) and 158bp spike (s) gene specific to SARS-COV-2 were detected by real-time reverse-transcription–polymerasechain-reaction (RT-PCR) [1], or if two targets (open reading frame 1a or 1b, nucleocapsid protein) were detected by RT-PCR [6]. Diagnostic criteria of COVID-19 of the Chinese Center for Disease Control and Prevention (CDC) is as follow. Suspected or probable cases are defined as cases that meet: (1) three clinical standards or (2) two clinical standards and one epidemiological criteria. Clinical criteria are: fever; radiographic evidence of pneumonia or acute respiratory distress syndrome; low or normal white blood cell count, or low lymphocyte count. Epidemiological criteria are: living in Wuhan or have a history of travel to Wuhan within 14 days before the onset of symptoms; contact with patients with fever and respiratory infection symptoms within 14 days before the onset of symptoms; and a link to any confirmed cases or clusters of suspected cases [11]. For the first case in a province, a confirmed case is defined as a suspected or probable case with positive viral nucleic acid detected at the municipal and provincial CDC. For the second case and all subsequent cases, it is defined as suspected or probable cases with positive viral nucleic acid detected at the municipal CDC [11].

□

SARS-COV-2 is a novel virus previously unknown. Numerous studies have revealed its genetic sequence and source. A team led by Professor Zhang YZ from Shanghai Public Health Clinical Center released the first SARS-COV-2 gene sequence on the website of viral.org on January 11, 2020 [12]. On January 12, 2020, five other virus genome sequences from different patients were released by the National Health Commission (NHC) of the People’s Republic of China in Global Initiative of Sharing All Influenza Data (GISAID), a global shared influenza virus database [13]. As of January 19, 2020, GISAID (http://gisaid.org/) has released 13 SARS-COV-2 genome sequences. One virus strain (Virus name: BetaCoV/Wuhan-Hu-1/2019; Accession ID: EPI_ISL_402125) has been released on GenBank (https://www.ncbi.nlm.nih.gov/nuccore/MN908947) by the Shanghai Public Health Clinical Center. Meanwhile, Chan JF et al. Constructed phylogenetic trees by genetic analysis of RdRp and S gene sequences of PCR amplified fragments from five patients, and revealed that SARS-COV-2 is closely related to bat SARS-like bat-SL-CoVZXC21 (NCBI accession number MG772934) and bat-SL-CoVZC45 (NCBI accession number MG772933) [1]. On January 23, 2020, a team led by Shi ZL form Wuhan Institute of Virology published an article revealed that genome of SARS-COV-2 has 96.2% similarity compared to that of RaTG13, a bat coronavirus previously detected on the Rhinolophus sinicus in Yunnan province and 79.5% similarity compared to that of SARS coronavirus [14]. And one study has suggested that the intermediate host of SARS-COV-2 may be snake [15].

It was suggested that human-to-human transmission may have occurred in Wuhan in the early stage of outbreak. Based on the data analysis of COVID-19 incidence in Wuhan from December 10, 2019 to January 4, 2020, it was shown that although most of the early COVID-19 had a history of Huanan Seafood Wholesale Market exposure, since the end of December 2019, the number of cases without a history of exposure had grown exponentially [6]. And from January 13, 2020 when Wuhan officially announced the outbreak of COVID-19 to January 21, 2020, the sudden decrease in the number of cases should be due to the underestimation of the number of cases and the delay in the confirmation report [6]. Several studies have estimated the transmission dynamics parameters of COVID-19. In the early stage of COVID-19 outbreak, basic reproductive number R_0_ was estimated to be 2.2 (95% confidence interval, 1.4 to 3.9) [6]. Of the 45 people who became ill before 1 January, the average duration from illness onset to first medical visit was 5.8 days (95% confidence interval, 4.3 to 7.5); of the 44 people who became ill before 1 January, the average interval from the disease onset to hospitalization was 12.5 days (95% confidence interval, 10.3 to 14.8) [6]. A study by the University of Hong Kong estimated that the basic reproductive number R_0_ was 2.68 and the doubling time of the number of cases was 6.4 days [11].

In the early stage of COVID-19 outbreak, the medical resources in Wuhan City were in short supply; the hospitals were overcrowded; the number of hospital beds and medical staff is insufficient; the corresponding diagnostic reagents were lacking; and the uniform diagnostic standards were missing. Some people don’t pay much attention to COVID-19, and some SARS-COV-2 infected patients with mild illness do not go to hospital. That all made the SARS-COV-2 infected cases could not be accurately diagnosed in time. Local administrators and health management officials were not aware of the severity of the SARS-COV-2 epidemic. They were afraid to take responsibility, so they may cover up the number of cases deliberately and report them late. The reporting rate may be significantly lower in Wuhan city in early stage due to previously described reasons. WHO Collaborating Centre for Infectious disease modelling estimated that the number of SARS-COV-2 infected cases in Wuhan by 18 January, 2020 was approximately 4000 (95% confidence interval: 1000-9700) [16]. The number is much larger than officially announced. Hiroshi Nishiura et al. through a study on the number of cases exported from Wuhan and the spatial back-calculation method, suggested that by January 24, 2020, the cumulative number of cases in China was about 5502 (95% confidence interval: 3027, 9057) [17]. As we all know, the emergence of super-spreaders is very important for the rapid spread of SARS-COV-2 in the early stage. In view of the event of one patient spread the virus to fourteen medical personnel, it is suggested that epidemic super-spreaders were generated in the early stage of virus transmission from animals to humans and in the early stage of transmission outbreak [3]. Therefore, in the early transmission stage of COVID-19, the number of cases reported by Wuhan officially should be lower than the actual number of cases in Wuhan.

In this study, we built SEIR models with a medical resource limitation factor that model the maximum number of people could be treated and isolated. We used a back propagation model to estimate the number of potential infections, and divided the epidemic area into three parts, Wuhan, Hubei (except Wuhan) and China (except Hubei) based on the different transmission pattern. In our study, we believed that COVID-19 has a R_0_ value of 3.6, the spread of SARS-COV-2 in China was effectively suppressed and the epidemic will end in early April.

According to the worldwide public reports of various media, Chinese People, governments at all levels have taken active measures to control the spread of COVID-19. According to our research, these measures effectively controlled the spread of COVID-19. The following measures have important reference significances for the control of infectious diseases may occur in the future. On December 31, 2019, the NHC expert group arrived in Wuhan to carry out the relevant testing and verification work. On January 01, 2020, Wuhan City shut down the Huanan Seafood Wholesale Market. On January 07, 2020, the NHC expert group initially identified the pathogen in these unexplained cases of viral pneumonia as a novel coronavirus. On January 20, 2020, Chinese government issued important instructions saying that the recent outbreak of pneumonia with novel coronavirus infection in Wuhan City, Hubei Province must be given great attention and all efforts to prevent and control. On January 21, 2020, The NHC issued 2020 No. 1 Proclamation to incorporate the novel coronavirus pneumonia into the Class B infectious diseases and take the prevention and control measures of Class A infectious diseases. At 10 a.m. on January 23, 2020, Wuhan City Bus, Subway, Ferry and Long-distance Passenger Transport was suspended, and the airport, railway station to leave Wuhan was temporarily closed. On January 24, 2020, Hubei Province launched the major public health emergency Level I response, and then as of 8 p.m. on January 25, 2020, a total of 30 provinces (regions, cities) in the country launched the major public health emergency I-level response. On January 27, 2020, the General Office of the State Council extended the Spring Festival holiday to February 2. On February 4, 2020, the NHC issued the “Guidelines for the Diagnosis and Treatment of Novel Coronavirus (2019-nCoV) Infection by the National Health Commission (Trial Version 5)”. On February 4, 2020, a designated makeshift infirmary named Wuhan Fire God Hill Hospital began to officially receive confirmed cases. On February 8, 2020, another designated makeshift infirmary Thunder God Hill Hospital was delivered to receive confirmed cases. On February 5, 2020, three “mobile field hospitals” began to be built in Wuhan. By February 8, 2020, central government of China has mobilized some 11000 medical personnel to support Wuhan. On February 9, 2020, Wuhan city took measures to try to collect all the suspected cases and confirmed cases of mild illness into the hospital for centralized treatment. On 9 February 2020, another group of medical personnel of some 6,000 people entered Wuhan to provide medical support.

Meanwhile, China’s scientific research institutions and medical equipment manufacturing enterprises quickly took measures to speed up scientific research and production of medical supplies. Scientific research institutions in China have done a lot of in-depth researches on the source, sequencing of SARS-COV-2, disease transmission dynamics, clinical manifestations, drug treatment, diagnostic standards, diagnostic reagents of COVID-19, etc. On January 26, 2020, China food and Drug Administration approved four SARS-COV-2 detection products of four companies to fully serve epidemic control needs. On February 7, 2020, Chinese SARS-COV-2 test kits daily production number exceeded 700,000, reaching the maximum production capacity of about 65%. By February 1, 2020, the daily production of medical protective clothing in China has been restored to 20000 pieces.

In the first stage of SARS-COV-2 transmission in Wuhan, the true number of infections was much higher than the officially announced probably because that people did not know COVID-19 is contagious and the local government did not take any control measure. In this stage, the infected cases were not effectively diagnosed, isolated, and treated, which led to rapid transmission of COVID-19. In the second stage, the trend of exponential growth of infection was suddenly controlled. The reason may be that Wuhan city was closed, people got the news that COVID-19 was contagious, and more and more medical resources were put into Wuhan. The medical capacity was still not sufficient to isolate every suspected or confirmed case and many infected people could not been treated or isolated timely. In the third stage, infected number growth curve in Wuhan showed a turning point and a gradual decline, possibly due to the input of medical resources and the strengthening of control measures. In the fourth stage, after February 13, 2020, the number of infected cases decreased rapidly, which is likely due to the substantial investment of medical resources and the strengthening of control measures, including the adoption of the new diagnostic standard “clinical diagnostic standard”. In the first stage of SARS-COV-2 transmission in Hubei (except Wuhan), the infected number increased exponentially should because of the infected people from Wuhan were not all quarantined and the medial resource were not sufficient. In this stage, the central government decided that one other province should support one city in Hubei. In the second stage, nearly all the infected and suspected people were quarantined, the added infected number reached the peak. The epidemic in China (except Hubei) was not so serious because of the sufficient medical resources and strong government control policy. In Stage one (from January 23, 2020 to February 3, 2020), the infected cases increased rapidly because of these virus carriers mainly infected his or her family members. In stage two (from February 4), most people canceled all their celebration activities and kept staying at home, and it greatly reduced the probability of infection. Anyone migrated from Hubei will be quarantined for at least 14 days. These dramatically reduced exposed number and the infected number got its peek around February 3, 2020.

The measures have effectively suppressed the spread of COVID-19. The infectious will spread sharply when R is greater than 1 and will gradually disappear when R is less than 1. Over time, R gradually decreased from 3.6 (Wuhan, stage 1), 3.4 (Hubei except Wuhan, stage 1) and 3.3 (China except Hubei, stage 1) to 0.67 (Wuhan, stage 4), 0.83 (Hubei except Wuhan, stage 2) and 0.63 (China except Hubei, stage 2), respectively. Especially after January 23, 2020 when Wuhan City was closed, the infected number showed a turning point in Wuhan in the next two weeks.

The outbreak of COVID-19 posed a major challenge to China’s Health system. Although the early human-to-human transmissions were described in the scientific literature, the local government of Hubei was not aware of the severity and did not inform the public timely, resulting in the public have no awareness of protection. Meanwhile, more than 5 million people left Wuhan with about 1960 infected among them during this time, and the outbreak spread across the country. At the very early stage of the outbreak of COVID-19, hospitals in Hubei Province had a serious shortage of medical protective equipment in routine reserves, which became the key limitation factor in controlling the outbreak. These defects should be improved in the future.

On February 12, 2020, the new diagnostic standard “clinical diagnostic standard” was adopted in Hubei province. Vast majority of new clinically diagnosed cases were diagnosed from accumulated suspected cases. Clinical diagnosis standard is different from the laboratory diagnosis standard widely used in the world. On February 12, 2020, Health Commission of Hubei Province announced that there were 14840 new confirmed cases in Hubei (including 13,332 clinically diagnosed cases). This does not mean that the epidemic trend has become worse. It is just a reflection of the change of diagnostic standards and the intensification of control measures. Data from Official of Hubei Province on February 12, 2020 were not used in this study, but could be used to evaluate some results of this study.

There were some limitations in this study, first the migrate data in the model was from Baidu Qianxi data set which might not cover all the population migrated. Second, there was no accurate medical resource limitation published especially in Wuhan and we estimated the media limitation by the maximum number of infected announced in the early stage and the added capacity of new hospitals in the later stage.

## Data Availability

All relevant data are within the manuscript.

## Acknowledgments

None

## Author Contributions

Conceptualization: Bo Zhang, Hongwei Zhou.

Data curation: Bo Zhang, Fang Zhou.

Formal analysis: Bo Zhang, Hongwei Zhou, Fang Zhou.

Methodology: Bo Zhang, Hongwei Zhou, Fang Zhou.

Supervision: Hongwei Zhou, Fang Zhou.

Validation: Hongwei Zhou, Fang Zhou.

Writing – original draft: Bo Zhang, Hongwei Zhou.

Writing – review & editing: Fang Zhou.

## Data Availability Statement

All relevant data are within the manuscript.

## References

1. Chan JF, Yuan S, Kok KH, To KK, Chu H, Yang J, et al. A familial cluster of pneumonia associated with the 2019 novel coronavirus indicating person-to-person transmission: a study of a family cluster. Lancet. 2020. https://doi.org/10.1016/S0140-6736(20)30154-9 PMID: 31986261

2. Martin Enserink. Update: ‘A bit chaotic.’ Christening of new coronavirus and its disease name create confusion. sciencemag. 2020 Feb 12 [Cited 2020 February 17]. Available from: https://www.sciencemag.org/news/2020/02/bit-chaotic-christening-new-coronavirus-and-its-disease-name-create-confusion

3. Li X, Zai J, Wang X, Li Y. Potential of large ‘first generation’ human-to-human transmission of 2019-nCoV. J Med Virol. 2020. https://doi.org/10.1002/jmv.25693 PMID: 31997390

4. Huang CL, Wang YM, Li XW, Ren LL, Zhao JP, Hu Y, et al. Clinical features of patients infected with 2019 novel coronavirus in Wuhan, China. Lancet. 2020. https://doi.org/10.1016/S0140-6736(20)30183-5 PMID: 31986264

5. Holshue ML, DeBolt C, Lindquist S, Lofy KH, Wiesman J, Bruce H, et al. First Case of 2019 Novel Coronavirus in the United States. N Engl J Med. 2020. https://doi.org/10.1056/NEJMoa2001191 PMID: 32004427

6. Li Q, Guan XH, Wu P, Wang XY, Zhou L, Tong YQ, et al. Early Transmission Dynamics in Wuhan, China, of Novel Coronavirus-Infected Pneumonia. N Engl J Med. 2020. https://doi.org/10.1056/NEJMoa2001316 PMID: 31995857

7. Rothe C, Schunk M, Sothmann P, Bretzel G, Froeschl G, Wallrauch C, et al. Transmission of 2019-nCoV Infection from an Asymptomatic Contact in Germany. N Engl J Med. 2020. https://doi.org/10.1056/NEJMc2001468 PMID: 32003551

8. Yoo JH. The Fight against the 2019-nCoV Outbreak: an Arduous March Has Just Begun. J Korean Med Sci. 2020; 35: e56. https://doi.org/10.3346/jkms.2020.35.e56 PMID: 31997618

9. Lipsitch M; Cohen T; Cooper B, Robins JM, Ma S, James L, et al. Transmission dynamics and control of severe acute respiratory syndrome. Science. 2003; 300: 1966–70. https://doi.org/10.1126/science.1086616 PMID: 12766207

10. Carlos WG, Dela Cruz CS; Cao B, Pasnick S, Jamil S. Novel Wuhan (2019-nCoV) Coronavirus. Am J Respir Crit Care Med. 2020. https://doi.org/10.1164/rccm.2014P7 PMID: 32004066

11. Wu JT; Leung K; Leung GM. Nowcasting and forecasting the potential domestic and international spread of the 2019-nCoV outbreak originating in Wuhan, China: a modelling study. Lancet. 2020. https://doi.org/10.1016/S0140-6736(20)30260-9 PMID: 32014114

12. Cohen J. Chinese researchers reveal draft genome of virus implicated in Wuhan pneumonia outbreak. American Association for the Advancement of Science. 2020 Jan 11 [Cited 2020 February 12]. Available from:https://www.sciencemag.org/news/2020/01/chinese-researchers-reveal-draft-genome-virus-implicated-wuhan-pneumonia-outbreak

13. Cohen J. Mining coronavirus genomes for clues to the outbreak’s origins [cited 2013 Oct 5]. Database: GISAID [Internet]. Available from: https://www.sciencemag.org/news/2020/01/mining-coronavirus-genomes-clues-outbreak-s-origins

14. Zhou P, Yang XL, Wang XG, Hu B, Zhang L, Zhang W, et al. Discovery of a novel coronavirus associated with the recent pneumonia outbreak in humans and its potential bat origin. bioRxiv. 2020. https://doi.org/:10.1101/2020.01.22.914952

15. Ji W, Wang W, Zhao XF, Zai JJ, Li XG. Homologous recombination within the spike glycoprotein of the newly identified coronavirus may boost cross-species transmission from snake to human. J Med Virol. 2020. https://doi.org/10.1002/jmv.25682 (2020) PMID: 31967321

16. Lu HZ. Drug treatment options for the 2019-new coronavirus (2019-nCoV). Biosci Trends. 2020. https://doi.org/10.5582/bst.2020.01020 PMID: 31996494

17. Nishiura H, Jung SM, Linton NM, Kinoshita R, Yang Y, Hayashi K, et al. The Extent of Transmission of Novel Coronavirus in Wuhan, China, 2020. J Clin Med. 2020. https://doi.org/10.3390/jcm9020330 PMID: 31991628

